# High seroprevalence of anti-SARS-CoV-2 antibodies after the first wave of the COVID-19 pandemic in a vulnerable population in Perpignan, France

**DOI:** 10.1101/2021.03.05.21252835

**Authors:** Adeline Beaumont, Cécile Durand, Martine Ledrans, Valérie Schwoebel, Harold Noel, Yann Le Strat, Donatien Diulius, Léa Colombain, Marie Médus, Philippe Gueudet, Damien Mouly, Hugues Aumaître

**Affiliations:** Santé Publique France (SpF), Direction des Régions, F-94415 Saint-Maurice, France; Santé Publique France (SpF), F-94415 Saint-Maurice, France; Agence Régionale de Santé (ARS) Occitanie, Délégation départementale des Pyrénées-Orientales, F-66020 Perpignan, France; Centre Hospitalier de Perpignan, Service des Maladies Infectieuses et Tropicales, F-66046 Perpignan, France; Centre Hospitalier de Perpignan, Laboratoire de biochimie et de biologie, F-66046 Perpignan, France

**Author notes:** Correspondence to Damien Mouly, Santé Publique France, Direction des Régions, F-94415 Saint-Maurice, France.

## Abstract

**Background:** In March 2020, many cases of COVID-19 were reported in three socially deprived neighbourhoods of the city of Perpignan, in the south of France, where large sedentary gypsy communities live. A study to measure seroprevalence was conducted in July 2020 to assess the level of contamination in these neighbourhoods after the first wave of the pandemic, and to identify factors associated with seropositivity.

**Methods:** SCoPe is a cross-sectional survey conducted in selected persons aged six years old and over living in three neighbourhoods in Perpignan. Households were selected by systematic sampling and participants by random sampling. Collected blood samples were tested for SARS-CoV-2 IgG and IgM antibodies using the EIecsys® immunoassay to target the coronavirus’s spike protein. Antibody seroprevalence was estimated from weighted data and associated factors were investigated using multivariate logistic regression.

**Results:** The seroprevalence of anti-SARS-CoV-2 antibodies was 35.4% (95% CI: 30.2-41.0). Over a fifth of seropositive individuals (21.7% ([14.1-31.8]) did not report any COVID-19 symptom. People aged 15-64 years old were at greater risk of seropositivity than those aged 65 years or over. Obesity prevalence was 40.7% (35.8-45.8) and obese people were more likely to be seropositive (aOR=2.0 [1.1-3.8]). The risk of being seropositive was higher in households with clinical COVID-19 cases (One case: aOR=2.5 [1.3-5.0]). In the neighbourhood with the highest measured seroprevalence, people living in a dwelling with 1-2 rooms had a higher risk of being seropositive than those living in a 4-room house (aOR=2.8 [1.2-6.3]). Working during the lockdown was associated with a lower risk of seropositivity (aOR=0.2 [0.03-1.0]).

**Conclusion:** Transmission prevalence of the SARS-COV-2 virus in this vulnerable population was very high during the COVID-19 pandemic’s first wave. Our results highlight the need to strengthen and adapt preventive measures by taking into account all social determinants of health, especially housing conditions.

## INTRODUCTION

With the emergence of COVID-19 and the resulting pandemic, questions about social inequalities in health during the current crisis have been raised^1^. Many health issues are involved, including inequalities in exposure to the SARS-CoV-2 virus, in the severity of the COVID-19 disease, and in access to healthcare^1, 2^. These concerns are all the more important given that these health inequalities are often cumulative^3^, leading to a marked risk of increased social deprivation in vulnerable populations^2,4^. Furthermore, lockdowns implemented in many countries have exacerbated pre-existing health inequalities.

During the ongoing epidemic, special attention has been given to the some 10,000 residents living in three of the poorest neighbourhoods (Haut-Vernet, Nouveau Logis and Saint-Jacques) in all of France. Located in the city of Perpignan (120,000 inhabitants, Occitania region), the employment rate is very low in these neighbourhoods, with only 25 to 30% of 15-64 year olds having work^5^. Sedentary gypsy communities make up a large part of the neighbourhoods’ population and share commonalities in lifestyle and culture, with the roles of family and religion being especially important. In Europe, gypsy communities have lower education levels and higher unemployment rates than the general public. They often have poorer living conditions and commonly face social exclusion^6^. Furthermore, their health literacy level is low. Their perception of health is that no illness exists if there are no visible signs^7^. Moreover, they have a poorer health status than that of the general population and face greater barriers to accessing healthcare^8-10^.

The first wave of the COVID-19 pandemic hit France at the beginning of 2020, leading to a national lockdown between 17 March and 11 May 2020. After the first positive case in Perpignan was detected using RT-PCR on 11 March 2020, the epidemic progressed rapidly in the city. On 20 March 2020, there were 47 confirmed cases in all the Pyrénées-Orientales ‘department’ (administrative area larger than a district but smaller than a region) (475,000 inhabitants) where Perpignan is located. On the same day, the intensive care unit in Perpignan hospital reported 19 people hospitalised and 5 deaths. An analysis by the hospital’s infectious and tropical diseases unit of all those diagnosed positive indicated that most of the patients were living in the three neighbourhoods described above. In order to control the situation, a curfew was implemented throughout the city beginning 21 March 2020 and accommodation facilities were offered to facilitate isolating positive cases and persons the latter had been in contact with. Outpatient medical centres were rapidly opened in the city’s most affected neighbourhoods to provide care to clinical cases and to prevent the spread of the virus in less impacted neighbourhoods. Specific surveillance based on data from these centres was also set up to monitor the evolution of the epidemic^11^. The mobilisation of various health and local actors ensured the swift dissemination of specific prevention information to the population throughout the first wave. On 1 May 2020, the epidemic had largely dissipated and two months after the lockdown, viral circulation was close to zero in Perpignan.

In this context, we conducted a seroprevalence study of anti-SARS-CoV-2 antibodies in Perpignan (SCoPe) in the three neighbourhoods described above to estimate the level of contamination during the first epidemic wave. In addition, we analysed environmental and behavioural factors in order to identify factors associated with increased viral circulation.

## METHODS

### Study design and participants

SCoPe is a cross-sectional seroprevalence survey of a sample of the population living in three neighbourhoods (Saint-Jacques (neighbourhood A), Haut-Vernet (neighbourhood B) and Nouveau Logis (neighbourhood C)) in the city of Perpignan (Figure 1). It was conducted between 29 June and 17 July 2020.

**Figure 1.**
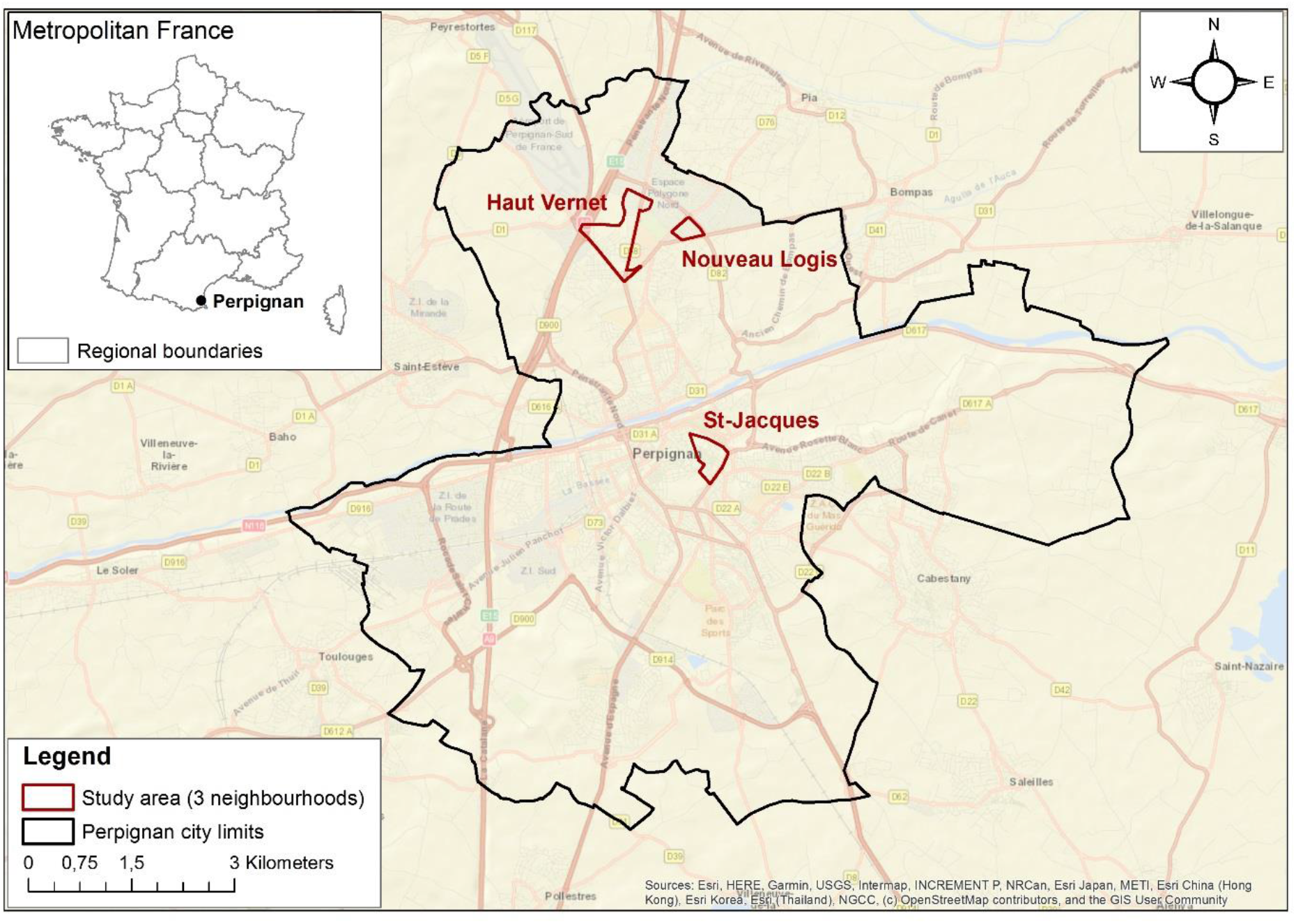
Map of the city of Perpignan and the three neighbourhoods studied

The limits of neighbourhoods A and B were demarcated using data from the French National Institute of Statistics and Economic Studies (INSEE), and neighbourhood C from city data (priority neighbourhood for social actions).

As sampling frames were unavailable for inhabitants or dwellings, we chose a two-stage random sampling process (households, inhabitants) stratified by neighbourhood. The field investigators criss-crossed each neighbourhood to select households for potential participation by systematic sampling from a predefined route and sampling interval generated by the research team. Depending on the household size, from one to four participants were then randomly recruited from households which agreed to participate (see: Supplemental materials - Survey procedure and logistics). Recruitment was carried out by teams of field investigators comprising members of the gypsy community and local social workers.

Individuals were eligible if they were 6 years old or over, had resided in the study area between 1 January 2020 and the survey date, were physically and mentally able to move to one of the study’s five purpose-built survey centres, and able to answer the survey questionnaire.

Participants were referred to the neighbourhood’s survey centre, where physicians used a standardized questionnaire in French - specifically designed for SCoPE - to collect information on the following: socio-demographic characteristics, medical conditions associated with the risk of severe COVID-19^12^, occurrence of symptoms suggestive of COVID-19 and healthcare seeking behaviour since 24 February 2020, characteristics of both the household and the housing the participant lived in during the first lockdown, knowledge of COVID-19 prevention measures, and behaviours during the first lockdown (see: Supplemental materials - Questionnaire). BMI was calculated by measuring height and weight. The questionnaire was designed in collaboration with local mediators in order to ensure that it would be acceptable to the study population and that they could understand it. A blood sample was collected by venepuncture for each participant: 3.5 ml for those aged 18 years old and over, and 600μl for those aged 6-17 years old.

The study protocol was approved by a French ethics committee (*Comité de Protection des Personnes Sud Est II*, Lyon, 2020-A01828-31). All participants were informed about the processing of personal data and of their rights. All gave their prior oral consent to participate. For those under 18 years of age, a parent or legal guardian provided consent.

### Laboratory analysis

The samples were sent to the laboratory at Perpignan hospital at room temperature (18-25°C) after a maximum storage time of 12 hours at maximum temperature of 5°C.

Serological tests were performed using EIecsys Anti-SARS-CoV-2^13^, an immunoassay for *in vitro* qualitative detection of immunoglobulin M (IgM) and immunoglobulin G (IgG) antibodies against the SARS-CoV-2 spike (S) protein in serum. Its sensitivity is 99.5% (97-100) at ≥ 14 days after PCR confirmation. Overall specificity is 99.8% (99.69-99.88)^13^.

### Statistical analysis

SCoPe’s estimations take into account the sampling design components (stages, sampling weights, stratification). Data were weighted by the inverse of the probability of selection (sampling weight) and adjusted for the age and sex in each neighbourhood from data of selected persons who declined to participate in the study, and from post-stratification using data from the most recent population census (2017).

A person was defined seropositive if anti-SARS-CoV-2 antibodies (IgM or IgG) were detected by the immunoassay. Seroprevalence (i.e., the proportion of seropositive individuals) was estimated with a 95% confidence interval (CI). It was compared between neighbourhoods and according to individual characteristics using the adjusted Wald F test. The association between seropositivity and reported symptoms was investigated in univariate analysis. Factors associated with seropositivity were then analysed using a multivariate logistic regression which took into account the sampling design. Behaviours during the lockdown were excluded from this analysis, except for leaving home to go to work. A forward selection procedure was applied with age, sex and neighbourhood being forced into the model. Variables with a p-value <0.1 were retained in the multivariate model and interactions were tested. A p-value <0.05 was considered statistically significant. Data were analysed using Stata V14.2 software (StataCorp, College Station, TX, USA).

## RESULTS

Of the total 1117 households initially selected for the study, 853 were visited and invited to participate (Figure 2). Of the latter, 628 (73.6%) households with 2101 eligible individuals agreed to partake in the random participant selection stage. The rate of those agreeing to partake in this stage varied between all three neighbourhoods: 78.7% in neighbourhood A, 48.7% in neighbourhood B and 98.9% in neighbourhood C. Among the 1248 individuals subsequently selected at random from the 2101 who were eligible, 700 (56.1%) went to the survey centres and were included in the analysis (i.e., study population): 312 from neighbourhood A (48.4%), 173 from neighbourhood B (70.0%) and 215 from neighbourhood C (60.4%).

**Figure 2.**
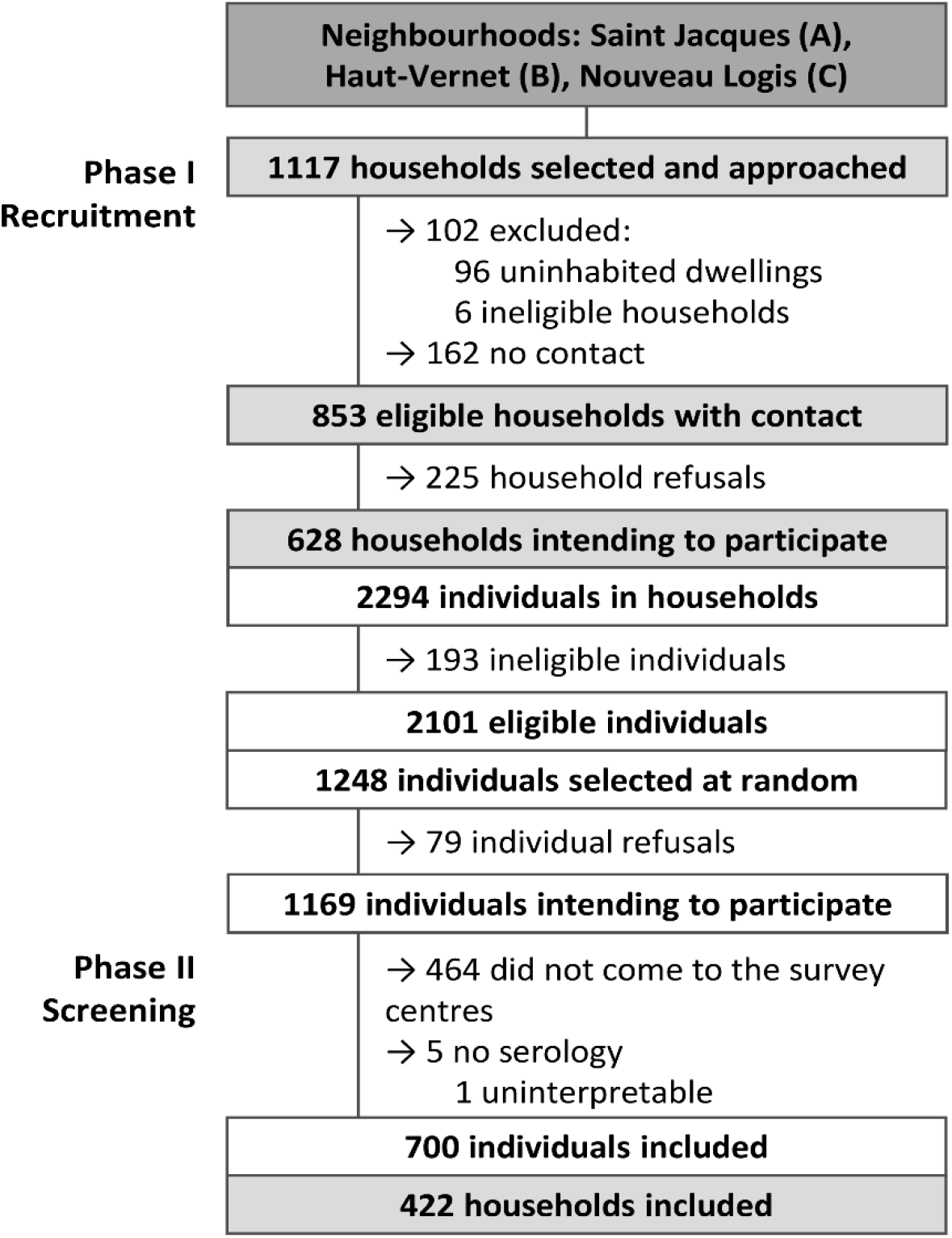
Flow chart of participants

### Study population

After weighting, females accounted for 50.4% of the study population. One third (34.3%) of the population was aged between 6 and 19 years old, 53.7% between 20 and 64 years old, while 12.0% were 65 years old or over.

Obesity prevalence was 40.7% (95% CI: 35.8-45.8): 43.5% (38.9-48.3) in adults (BMI≥30kg/m^2^) and 34.0% (22.2-48.2) in those aged 6-17 years old (BMI≥IOTF-30). Fifteen percent (13.0-17.3) of the study population reported having hypertension, 7.0% (5.5-8.8) heart disease, 9.4% (7.7-11.4) were being treated for diabetes, 5.5% (4.0-7.7) had asthma, while 4.9% (3.7-6.6) had (an)other chronic respiratory disease(s).

The majority of those in neighbourhood A were living in an apartment (71.5% [64.6-77.6]), while the majority of people in neighbourhoods B and C were living in a house (73.9% [62.8-82.6] and 83.9% [79.0-87.8], respectively). The number of people per room (except the living room) in each home was greater than one for 75.3% (69.9-80.1) of people living in neighbourhood A, for 55.5% (46.9-63.7) in neighbourhood B and for 80.5% (75.8-84.6) in neighbourhood C.

### Seroprevalence

Overall seroprevalence was estimated at 35.4% (30.2-41.0) for all three neighbourhoods. It was significantly higher in neighbourhood A (46.7% [39.0-54.7]) than in neighbourhoods B and C [13.9% [8.2-22.6] and 17.1% [13.0-22.2], respectively).

### Symptoms during the study period

Among seropositive people, 21.7% (14.1-31.8) reported no symptoms suggestive of COVID-19 during the study period (from 24 February 2020 to the survey date). One in seven (14.6% [9.5-21.9]) of those who reported no symptoms were tested seropositive. Seropositive people mostly reported unusual fatigue (58.9% [48.9-68.2]), a headache (51.7% [42.4-60.9]), ageusia/anosmia (49.8% [40.2-59.4]), a fever or a feeling of having a fever (49.1% [40.6-57.6]), a cough (46.4% [37.5-55.5]) and myalgia (45.7% [37.4-54.3]).

There was a significant positive association between seropositivity and symptoms (Odds Ratio (OR)=8.1 [4.5-14.6], p<0.001). Ageusia/anosmia were the symptoms most strongly associated with seropositivity (OR=14.8 [7.9-27.7], p<0.001), with positive and negative predictive values of 81.3% [71.5-88.3] and 77.3% [71.4-82.4], respectively. All other symptoms were also significantly associated with seropositivity, except for rhinorrhea (Figure 3).

**Figure 3.**
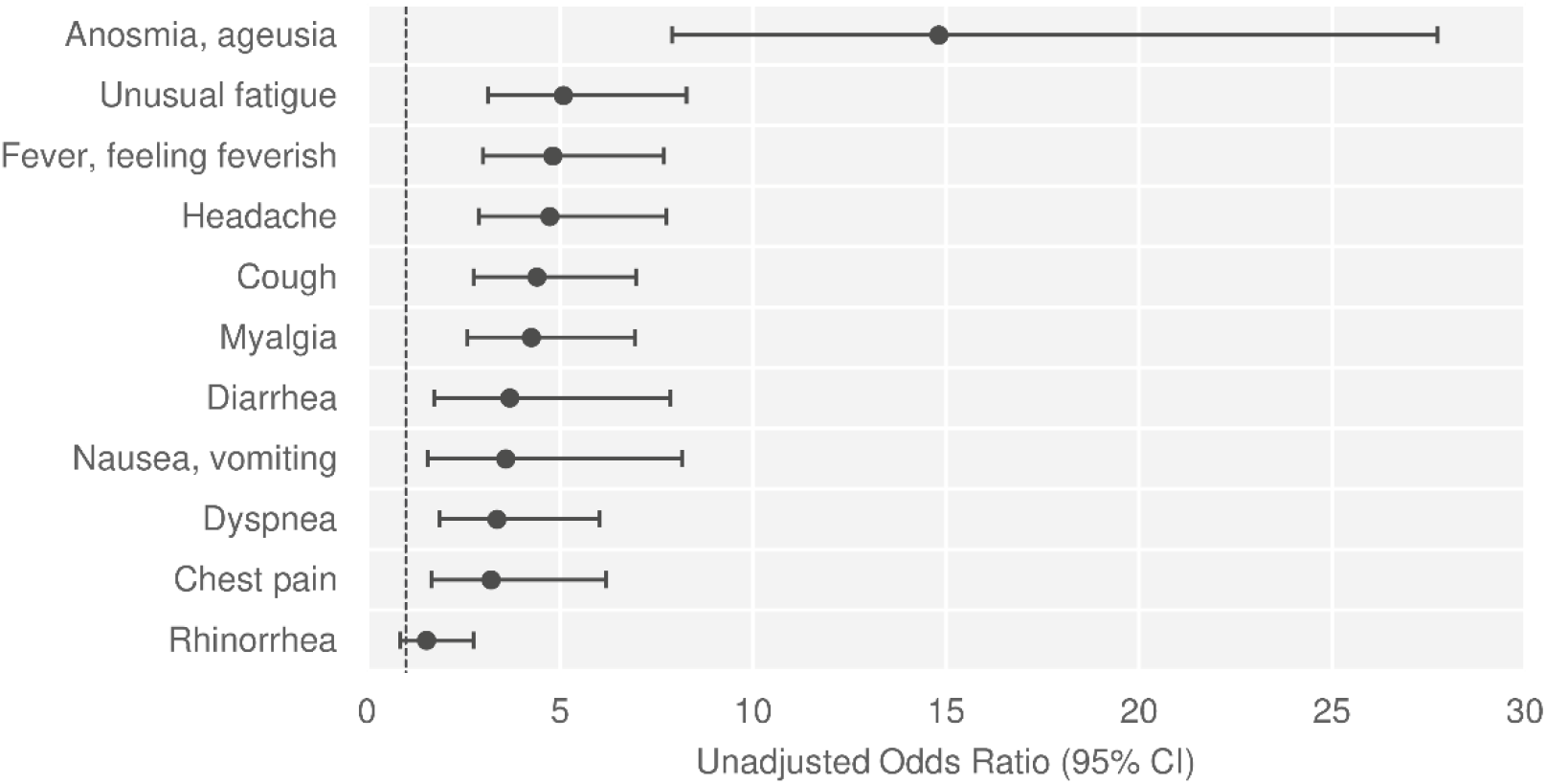
Association between seropositivity and reporting symptoms * Analysis performed on all sampled individuals (n=700) using simple logistic regressions.

### Healthcare seeking behaviours during the study period

During the study period, 15.8% (11.3-21.6) of symptomatic people consulted a COVID-19 centre when symptoms occurred and 9.6% (6.6-13.6) had a RT-PCR test (positive PCR=29.0%). Specifically, 41.8% of seropositive participants had had a positive RT-PCR test result.

Among seropositive participants, 7.9% (4.6-13.2) had been hospitalised during the study period, almost all having had medical conditions associated with severe COVID-19 (89.3%).

### Factors associated with seropositivity

In the univariate analysis (Table 1), people aged 65 years or over were less likely to be seropositive (p<0.001). No significant difference was observed between males and females regarding the likelihood of being seropositive. Obese people were more likely to be seropositive (OR=2.0 IC95%=[1.3-3.2], p=0.002). The presence of one (OR=3.0 [1.8-5.2], p<0.001) or more (OR=7.8 [4.0-15.2], p<0.001) clinical COVID-19 cases in the household was associated with a greater risk of seropositivity. People living in a dwelling with three or fewer rooms (1-2 rooms: OR=2.1 [1.2-3.8], p=0.011; 3 rooms: OR=2.2 [1.3-3.9], p=0.005) were more likely to be seropositive. The proportion of seropositive people increased with the number of people per room in the dwelling (p=0.001). People who worked during the lockdown were less likely to be seropositive (OR=0.1 [0.02-0.5], p=0.006). Furthermore, people who reported leaving their home once a week or less for walks during the lockdown were less likely to be seropositive than people who went out every day or almost every day (OR=0.2 [0.1-0.7], p=0.012).

**Table 1.**
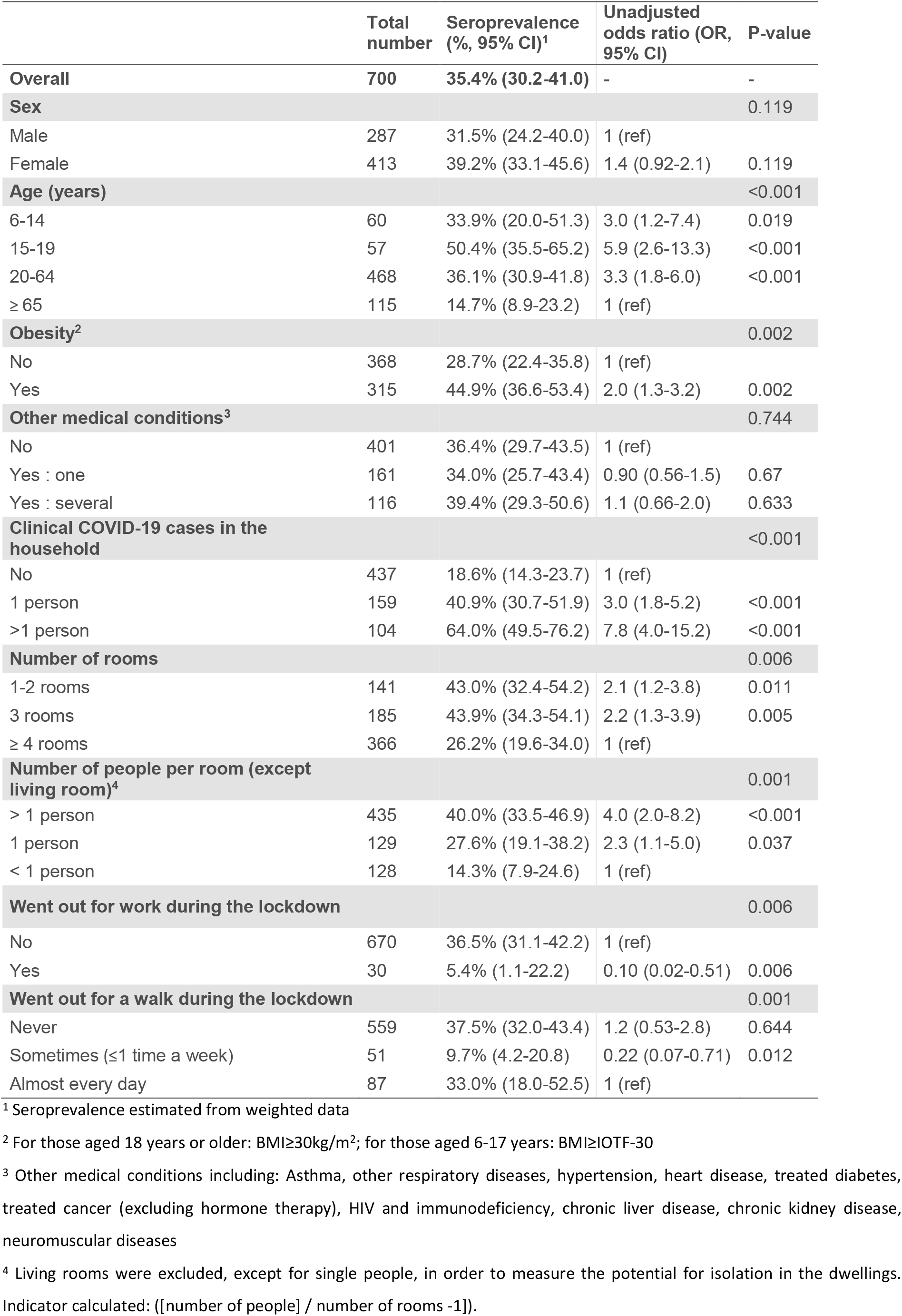
Factors associated with SARS-CoV-2 seropositivity: univariate analysis

|  | Total number | Seroprevalence (%; 95% CI)<sup>1</sup> | Unadjusted odds ratio (OR, 95% CI) | P-value |
| --- | --- | --- | --- | --- |
| **Overall** | **700** | **35.4% (30.2-41.0)** | **-** | **-** |
| **Sex** |  |  |  | 0.119 |
| Male | 287 | 31.5% (24.2-40.0) | 1 (ref) |  |
| Female | 413 | 39.2% (33.1-45.6) | 1.4 (0.92-2.1) | 0.119 |
| **Age (years)** |  |  |  | <0.001 |
| 6-14 | 60 | 33.9% (20.0-51.3) | 3.0 (1.2-7.4) | 0.019 |
| 15-19 | 57 | 50.4% (35.5-65.2) | 5.9 (2.6-13.3) | <0.001 |
| 20-64 | 468 | 36.1% (30.9-41.8) | 3.3 (1.8-6.0) | <0.001 |
| ≥ 65 | 115 | 14.7% (8.9-23.2) | 1 (ref) |  |
| **Obesity<sup>2</sup>** |  |  |  | 0.002 |
| No | 368 | 28.7% (22.4-35.8) | 1 (ref) |  |
| Yes | 315 | 44.9% (36.6-53.4) | 2.0 (1.3-3.2) | 0.002 |
| **Other medical conditions<sup>3</sup>** |  |  |  | 0.744 |
| No | 401 | 36.4% (29.7-43.5) | 1 (ref) |  |
| Yes : one | 161 | 34.0% (25.7-43.4) | 0.90 (0.56-1.5) | 0.67 |
| Yes : several | 116 | 39.4% (29.3-50.6) | 1.1 (0.66-2.0) | 0.633 |
| **Clinical COVID-19 cases in the household** |  |  |  | <0.001 |
| No | 437 | 18.6% (14.3-23.7) | 1 (ref) |  |
| 1 person | 159 | 40.9% (30.7-51.9) | 3.0 (1.8-5.2) | <0.001 |
| >1 person | 104 | 64.0% (49.5-76.2) | 7.8 (4.0-15.2) | <0.001 |
| **Number of rooms** |  |  |  | 0.006 |
| 1-2 rooms | 141 | 43.0% (32.4-54.2) | 2.1 (1.2-3.8) | 0.011 |
| 3 rooms | 185 | 43.9% (34.3-54.1) | 2.2 (1.3-3.9) | 0.005 |
| ≥ 4 rooms | 366 | 26.2% (19.6-34.0) | 1 (ref) |  |
| **Number of people per room (except living room)<sup>4</sup>** |  |  |  | 0.001 |
| > 1 person | 435 | 40.0% (33.5-46.9) | 4.0 (2.0-8.2) | <0.001 |
| 1 person | 129 | 27.6% (19.1-38.2) | 2.3 (1.1-5.0) | 0.037 |
| < 1 person | 128 | 14.3% (7.9-24.6) | 1 (ref) |  |
| **Went out for work during the lockdown** |  |  |  | 0.006 |
| No | 670 | 36.5% (31.1-42.2) | 1 (ref) |  |
| Yes | 30 | 5.4% (1.1-22.2) | 0.10 (0.02-0.51) | 0.006 |
| **Went out for a walk during the lockdown** |  |  |  | 0.001 |
| Never | 559 | 37.5% (32.0-43.4) | 1.2 (0.53-2.8) | 0.644 |
| Sometimes (≤1 time a week) | 51 | 9.7% (4.2-20.8) | 0.22 (0.07-0.71) | 0.012 |
| Almost every day | 87 | 33.0% (18.0-52.5) | 1 (ref) |  |

<sup>1</sup> Seroprevalence estimated from weighted data
<sup>2</sup> For those aged 18 years or older: BMI≥30kg/m<sup>2</sup>; for those aged 6-17 years: BMI≥IOTF-30
<sup>3</sup> Other medical conditions including: Asthma, other respiratory diseases, hypertension, heart disease, treated diabetes, treated cancer (excluding hormone therapy), HIV and immunodeficiency, chronic liver disease, chronic kidney disease, neuromuscular diseases
<sup>4</sup> Living rooms were excluded, except for single people, in order to measure the potential for isolation in the dwellings. Indicator calculated: ([number of people] / number of rooms -1)).

In the multivariate analysis (Table 2), the association between seropositivity and the presence of clinical cases in the household remained strong after adjusting for other factors (one person: adjusted odds ratio (aOR)=2.5 [1.3-5.0], p=0.007; ≥ 2 persons: aOR=6.9 [3.1-15.2], p<0.001). People aged 15-19 years (aOR 9.1 [2.8-29.8], p<0.001) and 20-64 years (aOR=4.5 (2.0-10.1), p<0.001) had a higher risk of being seropositive than those aged 65 years or over. Females were more likely to be seropositive than males (aOR=1.8 [1.0-3.3], p=0.034). Seropositivity was significantly associated with obesity (aOR=2.0 [1.1-3.8], p=0.02) and other medical conditions (aOR=3.2 [1.6-6.3], p=0.001). There was a significant interaction between the neighbourhood and the number of rooms in the dwelling (p=0.004). People living in a one- or two-room dwelling in neighbourhood A were more likely to be seropositive than those living in a dwelling with four or more rooms (aOR=2.8 [1.2-6.3]). Working during lockdown remained independently associated with decreased seropositivity (aOR=0.2 [0.03-1.0], p=0.05).

**Table 2.** Factors associated with SARS-CoV-2 seropositivity: multivariate analysis

|  | Adjusted Odds Ratio (aOR, 95% CI) | P-value |
| --- | --- | --- |
| **Sex** |  | 0.034 |
| Male | 1 (ref) |  |
| Female | 1.8 (1.0-3.3) | 0.034 |
| **Age (years)** |  | <0.001 |
| 6-14 | 1.8 (0.53-6.1) | 0.344 |
| 15-19 | 9.1 (2.8-29.8) | <0.001 |
| 20-64 | 4.5 (2.0-10.1) | <0.001 |
| ≥ 65 | 1 (ref) |  |
| **Obesity** |  | 0.024 |
| No | 1 (ref) |  |
| Yes | 2.0 (1.1-3.8) | 0.024 |
| **Other medical conditions** |  | 0.004 |
| No | 1 (ref) |  |
| Yes : one | 1.1 (0.57-2.0) | 0.863 |
| Yes : several | 3.2 (1.6-6.3) | 0.001 |
| **Clinical COVID-19 cases in the household** |  | <0.001 |
| No | 1 (ref) |  |
| 1 person | 2.5 (1.3-5.0) | 0.007 |
| >1 person | 6.9 (3.1-15.2) | <0.001 |
| **Went out for work during the lockdown** |  | 0.048 |
| No | 1 (ref) |  |
| Yes | 0.18 (0.03-1.0) | 0.048 |
| **Number of rooms by neighbourhood<sup>1</sup>** |  | 0.007 |
| **Neighbourhood A** |  |  |
| 1-2 | 2.8 (1.2-6.3) | 0.016 |
| 3 | 2.2 (1.0-5.0) | 0.064 |
| ≥ 4 | 1 (ref) |  |
| **Neighbourhood B** |  |  |
| 1-2 | 1.5 (0.3-6.4) | 0.594 |
| 3 | 0.23 (0.04-1.2) | 0.075 |
| ≥ 4 | 1 (ref) |  |
| **Neighbourhood C** |  |  |
| 1-2 | 0.58 (0.22-1.5) | 0.262 |
| 3 | 2.3 (0.91-5.9) | 0.078 |
| ≥ 4 | 1 (ref) |  |

<sup>1</sup> Model includes an interaction term: number of rooms\*neighbourhood

## DISCUSSION

Our findings from the SCoPe seroprevalence study in three socially deprived neighbourhoods with a large sedentary gypsy community in Perpignan indicate that more than one in three (35.4%) people developed antibodies against SARS-CoV-2 during the first months of the COVID-19 epidemic. In comparison, estimates for the general population in May 2020 indicated an antibody prevalence of 1.9% in the Occitania region (where Perpignan is situated) and less than 5% in France and Spain (Perpignan is located very close to the Spanish border)^14, 15^.

Although the proportion of asymptomatic SARS-CoV-2 infections varies greatly from one study to another, the proportion we found (21.7%) was comparable with the results of two meta-analyses (20% [17-25])^16^ (17% [14-20])^17^. The strong specificity of ageusia/anosmia symptoms has already been observed in other studies^18^. This specificity could be explored in greater depth in the context of developing a strategy for early diagnosis of COVID-19 and self-isolation.

Lower seroprevalence was reported among study participants aged 65 years and over. This may partially be explained by a result from a qualitative study simultaneously conducted with SCoPe^19^ which found that this older population went outdoors less frequently and had fewer social contacts during the first wave of the epidemic thanks to the very protective stance adopted by the local community. In addition, females were more likely to be seropositive in the multivariate analysis. The associations between seropositivity and age and between seropositivity and sex differ between studies, although several have found a lower seroprevalence among older people, particularly in France^14, 20^. The fact that few seroprevalence studies have been conducted to date in a similar context (high level of infection, socially deprived neighbourhood) could explain these differences.

Our results showed that obese people had higher seroprevalence of SARS-CoV-2 antibodies independently from other factors. This is consistent with the findings of a meta-analysis of 20 published studies on the subject (OR=1.46 [1.30-1.65])^21^. Obesity has been associated with low socioeconomic status^22^. The association we found between obesity and seropositivity may be explained by potential confounders linked to unfavourable socioeconomic conditions. SCoPe did not comprehensively measure these conditions for reasons of study acceptability. Metabolic and immune dysfunction and inflammatory mechanisms may be implicated in the clinical aggravation of COVID-19 in obese people^23, 24^. These mechanisms might also be involved in increasing the risk of infection, although this association is less well established. Prolonged viral shedding in obese people, something already seen for influenza^25^, may also occur for SARS-CoV-2 and could play a role in the spread of the virus in families where obesity is prevalent.

Our study also confirms findings elsewhere that the risk of transmission is greater when a clinical case is present in the same household^14, 15^. Working outside the home during the first lockdown was associated with a lower risk of seropositivity. This result may reflect a higher socioeconomic status of people who worked. It might also be explained by a reduction in close indoor contacts with other household members, something highlighted in a seroprevalence study among socially deprived populations living in overcrowded residences in the Paris region^26^.

In our study, seroprevalence was higher for people living in crowded housing, and after adjusting for other factors, small dwelling size was a significant associated factor, but only in neighbourhood A. This result was also found in other French studies^14, 26^. In addition, living conditions - not analysed in our study - may also explain the higher seroprevalence in this particular neighbourhood. Population density, a factor associated with higher seroprevalence elsewhere^14, 15^, was higher in neighbourhood A than in both other neighbourhoods. The majority of accommodation in neighbourhood A comprises flats, and almost one-quarter of all dwellings are less than 40m^2^ ^27^. Insalubrity was also very present in neighbourhood A, which is one of the priority areas in an ongoing national urban renewal programme^28^. Accordingly, ventilation problems, lack of outdoor space and overcrowding may explain the higher risk of contamination. In general, the community-based lifestyle of the gypsy population may also have increased the risk of contact with a clinical COVID-19 case.

Overall, we achieved a 56% participation rate in this difficult-to-reach population thanks to local mediators and contacts, whose collaboration was essential. Furthermore, despite the unavailability of sampling frames, the study was designed and implemented very quickly after the first wave ended, thanks to careful training and supervision of the interviewers throughout the field survey. This speed of implementation was necessary given the uncertainties surrounding the duration of SARS-CoV-2 IgG antibodies after infection.

Our study has several limitations. First, it was conducted 4 months after the first wave ended, leading to possible recall bias in the reporting of symptoms. The assessment of behaviours during lockdown was very complex because of the fact that their evolution was not measured during the course of the first wave. It is important to underline that a qualitative study observed a shift in the three neighbourhoods’ awareness of the dangers of COVID-19 following the first deaths, particularly that of a young woman. The same study observed a substantial improvement in compliance with prevention measures during the lockdown^19^. This is why the association between these behaviours and seropositivity (except for going out to work) was not studied in our analysis. Second, the systematic sampling method used to select households made it difficult to estimate the total number of individuals to approach. Third, we also had difficulties reaching some of the selected households, despite flyers being placed in letterboxes and several visits. Finally, selection bias may have occurred. More specifically, people with a history of COVID-19 type symptoms may have been more willing to participate in the study than people with no such history. It is also possible that people who had been tested positive before the study were less willing to participate. Incomplete data on reasons for non-response prevented us from further exploring this issue.

The high estimated seroprevalence after the first wave of SARS-CoV-2 infection in the three socially deprived neighbourhoods in the present study confirms the very high vulnerability to COVID-19 of populations living in socially deprived conditions, and underlines the need for more sophisticated surveillance and specific disease prevention measures^29^. Additional observations using a sociological approach, should provide an accurate assessment of the ability of this population to improve their level of health literacy and to assimilate protective measures. Although underlying mechanisms remain unclear, our results support previous findings that obese individuals are at higher risk of SARS-CoV-2 infection, and confirm the importance of conducting preventive interventions in this population. This is especially relevant as future vaccines might be less effective for these people^23, 25^. All future vaccination strategies should be designed to ensure that they are acceptable to this vulnerable population^30^.

The long-term protection of vulnerable populations such as that in the present study who are particularly exposed to health and environmental crises, must be improved by strengthening specific prevention and health promotion programmes and reducing social inequalities in health^31^. In this context, policies against substandard housing have a key role in improving living conditions. Finally, health strategies can only be successful by ensuring long-term partnerships with organisations and stakeholders capable of rapid mobilisation in the event of a crisis.

## Data Availability

Data may be obtained from a third party and are not publicly available: Anonymised data are available for researchers from the corresponding author, Damien Mouly, on reasonable request.

## ACKNOWLEDGMENTS

We would like to thank the field-based interviewers, the nursing staff (Centre Hospitalier de Perpignan, Perpignan) and Anne Guinard (Santé Publique France, Toulouse) who participated in the recruitment phase of the study. We thank Clothilde Hachin (Santé Publique France, Saint-Maurice) for her support on the study protocol, Séverine Bailleul (Santé Publique France, Toulouse) for her help during preparation of the study, and Gabrielle Jones (Santé Publique France, Saint-Maurice) for her technical support.

## REFERENCES

1. Bambra C, Riordan R, Ford J, et al. The COVID-19 pandemic and health inequalities. J Epidemiol Community Health 2020;74(11):964–68. doi: 10.1136/jech-2020-214401

2. DREES. Les inégalités sociales face à l’épidémie de Covid-19. Etat des lieux et perspectives [in French]. Les dossiers de la DREES 2020 July:40.

3. Bajos N, Warszawski J, Pailhé A, et al. Les inégalités sociales au temps du COVID-19 [in French].Questions de Santé Publique 2020 Oct:12.

4. Abedi V, Olulana O, Avula V, et al. Racial, Economic, and Health Inequality and COVID-19 Infection in the United States. J Racial Ethn Health Disparities 2020:1–11. doi: 10.1007/s40615-020-00833-4 [published Online First: 2020/09/03]

5. Bordet C, Bourniquel C, Flachère M, et al. Quartiers prioritaires de la politique de la ville en Occitanie : les multiples visages de la pauvreté. Insee Dossier Occitanie 2018 July:151–55.

6. Parekh N, Rose T. Health inequalities of the Roma in Europe: a literature review. Cent Eur J Public Health 2011;19(3):139–42. [published Online First: 2011/10/27]

7. Ramos-Morcillo AJ, Leal-Costa C, Hueso-Montoro C, et al. Concept of Health and Sickness of the Spanish Gypsy Population: A Qualitative Approach. Int J Environ Res Public Health 2019;16(22) doi: 10.3390/ijerph16224492 [published Online First: 2019/11/20]

8. Hajioff S, McKee M. The health of the Roma people: a review of the published literature. J Epidemiol Community Health 2000;54(11):864–9. doi: 10.1136/jech.54.11.864 [published Online First: 2000/10/12]

9. Cook B, Wayne GF, Valentine A, et al. Revisiting the evidence on health and health care disparities among the Roma: a systematic review 2003-2012. Int J Public Health 2013;58(6):885–911. doi: 10.1007/s00038-013-0518-6 [published Online First: 2013/10/08]

10. McFadden A, Siebelt L, Gavine A, et al. Gypsy, Roma and Traveller access to and engagement with health services: a systematic review. Eur J Public Health 2018;28(1):74–81. doi: 10.1093/eurpub/ckx226 [published Online First: 2018/01/19]

11. Simac L, Ledrans M, Catelinois O, et al. COVID-19 in the vulnerable population of the Saint-Jacques and Haut-Vernet districts of Perpignan (France): Health surveillance carried out using local data [in French]. Bull Epidémiol Hebd 2020(30):590–98.

12. Haut Conseil de la santé publique. Avis du 29 octobre 2020 relatif à l’actualisation de la liste des facteurs de risque de forme grave de Covid-19 [in French]. Paris, 2020:b34.

13. Muench P, Jochum S, Wenderoth V, et al. Development and Validation of the Elecsys Anti-SARS-CoV-2 Immunoassay as a Highly Specific Tool for Determining Past Exposure to SARS-CoV-2. J Clin Microbiol 2020;58(10) doi: 10.1128/JCM.01694-20 [published Online First: 2020/08/05]

14. Warszawski J, Bajos N, Meyer L, et al. En mai 2020, 4,5 % de la population en France métropolitaine a développé des anticorps contre le SARS-CoV-2 : premiers résultats de l’enquête nationale EpiCov [in French]. Études et résultats 2020 Oct:6.

15. Pollán M, Pérez-Gómez B, Pastor-Barriuso R, et al. Prevalence of SARS-CoV-2 in Spain (ENE-COVID): a nationwide, population-based seroepidemiological study. Lancet 2020;396(10250):535–44. doi: 10.1016/s0140-6736(20)31483-5 [published Online First: 2020/07/10]

16. Buitrago-Garcia D, Egli-Gany D, Counotte MJ, et al. Occurrence and transmission potential of asymptomatic and presymptomatic SARS-CoV-2 infections: A living systematic review and meta-analysis. PLoS Med 2020;17(9):e1003346. doi: 10.1371/journal.pmed.1003346 [published Online First: 2020/09/23]

17. Byambasuren O, Cardona M, Bell K, et al. Estimating the extent of asymptomatic COVID-19 and its potential for community transmission: Systematic review and meta-analysis. Official Journal of the Association of Medical Microbiology and Infectious Disease Canada 2020 doi: 10.3138/jammi-2020-0030

18. Makaronidis J, Mok J, Balogun N, et al. Seroprevalence of SARS-CoV-2 antibodies in people with an acute loss in their sense of smell and/or taste in a community-based population in London, UK: An observational cohort study. PLoS Med 2020;17(10):e1003358. doi: 10.1371/journal.pmed.1003358 [published Online First: 2020/10/02]

19. Srocynski M., Yeghicheyan. Les Gitans de Perpignan face à la Covid-19. Approche qualitative d’un cluster [in French]. Unpublished report. CREAI-ORS Occitanie; 2021 March.

20. Stringhini S, Wisniak A, Piumatti G, et al. Seroprevalence of anti-SARS-CoV-2 IgG antibodies in Geneva, Switzerland (SEROCoV-POP): a population-based study. Lancet 2020;396(10247):313–19. doi: 10.1016/S0140-6736(20)31304-0 [published Online First: 2020/06/15]

21. Popkin BM, D.S, Green WD, et al. Individuals with obesity and COVID-19: A global perspective on the epidemiology and biological relationships. Obes Rev 2020;21(11):e13128. doi: 10.1111/obr.13128 [published Online First: 2020/08/28]

22. Vernay M, Malon A, Oleko A, et al. Association of socioeconomic status with overall overweight and central obesity in men and women: the French Nutrition and Health Survey 2006. BMC Public Health 2009;9:215. doi: 10.1186/1471-2458-9-215 [published Online First: 2009/07/04]

23. Alberca RW, Oliveira LM, Branco A, et al. Obesity as a risk factor for COVID-19: an overview. Crit Rev Food Sci Nutr 2020:1–15. doi: 10.1080/10408398.2020.1775546 [published Online First: 2020/06/17]

24. Goossens GH, Dicker D, Farpour-Lambert NJ, et al. Obesity and COVID-19: A Perspective from the European Association for the Study of Obesity on Immunological Perturbations, Therapeutic Challenges, and Opportunities in Obesity. Obes Facts 2020;13(4):439–52. doi: 10.1159/000510719 [published Online First: 2020/08/14]

25. Luzi L, Radaelli MG. Influenza and obesity: its odd relationship and the lessons for COVID-19 pandemic. Acta Diabetol 2020;57(6):759–64. doi: 10.1007/s00592-020-01522-8 [published Online First: 2020/04/07]

26. Roederer T, Mollo B, Vincent C, et al. Seroprevalence and risk factors of exposure to COVID-19 in homeless people in Paris, France: a cross-sectional study. Lancet Public Health 2021 doi: 10.1016/s2468-2667(21)00001-3 [published Online First: 2021/02/09]

27. Logement en 2017 : Recensement de la population - Base infracommunale (IRIS) [in French] [Internet]. Insee. 2020 [cited 2020 Dec]. Available from: https://www.insee.fr/.

28. Arrêté du 29 avril 2015 relatif à la liste des quartiers prioritaires de la politique de la ville présentant les dysfonctionnements urbains les plus importants et visés en priorité par le nouveau programme national de renouvellement urbain [in French]: JORF n°0106 [NOR: VJSV1508731A]; 2015 [updated 2015 May 8; cited 2021 Jan 20]. Available from: https://www.legifrance.gouv.fr/loda/id/LEGITEXT000030556555/2020-08-28/ accessed Jan 20 2021.

29. Burström B, Tao W. Social determinants of health and inequalities in COVID-19. Eur J Public Health 2020;30(4):617–18. doi: 10.1093/eurpub/ckaa095 [published Online First: 2020/07/09]

30. Jackson C, Bedford H, Cheater FM, et al. Needles, Jabs and Jags: a qualitative exploration of barriers and facilitators to child and adult immunisation uptake among Gypsies, Travellers and Roma. BMC Public Health 2017;17(1):254. doi: 10.1186/s12889-017-4178-y [published Online First: 2017/03/16]

31. Fernández-Feito A, Pesquera-Cabezas R, González-Cobo C, et al. What do we know about the health of Spanish Roma people and what has been done to improve it? A scoping review. Ethn Health 2019;24(2):224–43. doi: 10.1080/13557858.2017.1315373 [published Online First: 2017/04/12]

