## supplemental_materials for "High seroprevalence of anti-SARS-CoV-2 antibodies after the first wave of the COVID-19 pandemic in a vulnerable population in Perpignan, France"

#### Survey procedure and logistics

All the interviewers were initially trained in the study method and the use of the survey tools (online questionnaire, household forms, logbooks). They were supervised throughout the survey. In total, the study mobilised almost 80 people.

Local mediators and social workers implemented the selection phase (households and inhabitants) and provided potential participants with information about the survey. For this purpose, they had a map of each neighbourhood describing the starting point and a specific route to be followed. These starting points were defined randomly from a grid of neighbourhood maps as follows: randomly drawing a grid cell and, after assigning a number to each building, randomly choosing one building on that grid cell to be the starting point. The starting direction was also randomly chosen. In neighbourhood A, every 2 out of 5 households were selected, while in neighbourhood B, every 2 out of 3 were selected. In neighbourhood C, all households were selected. These sampling intervals were determined according to the size of the targeted sample, the estimated non-response rate, the number of dwellings in the neighbourhood, and the average number of people per household in the population census. The sampling intervals were then adjusted to take into account field observations during the first days of the survey. The a priori targeted sample size was 1,000 participants, equally distributed among the three neighbourhoods, for an expected prevalence of 10%, a margin of error of 2.5 percentage points and a design effect of 2.

In each selected household, between one and four participants were randomly selected in proportion to the size of the household. The selected inhabitants received information notes and a ticket indicating their name and surname and the fact that they had been selected for participation. They were then invited to visit one of the five centres specifically set up for the survey (2 each in neighbourhoods A and B, 1 in neighbourhood C), and to bring their ticket with them to facilitate their identification and inclusion. All the initially selected households and their inhabitants were monitored using logbooks and forms to record household data. To increase the participation rate, three visits were made at different days and times. When no one was at home, a letter with a phone number was put in the letterbox. People who agreed to participate but who did not visit a survey centre were called back by phone.

In the survey centres, doctors and nurses from the infectious and tropical diseases unit (SMIT) of Perpignan hospital checked the identity of participants, administered a face-to-face questionnaire and took blood samples. Answers to the questionnaire items were entered in real-time online on a secure Voozanoo$^{TM}$ platform. Children under 18 years had to be accompanied by a parent or legal representative, and participants who did not speak French very well could - if they wished - be accompanied by a family member or friend who did. The weight and height of the participants were measured at the time of the questionnaire. A doctor gave individual results to those tested in July/August 2020.

### Questionnaire

French version / *English translation*

Date d'enquête :/ *Survey date:* …./…./2020

Centre de prélèvement : / *Test centre*

- ☐ St Jacques
- ☐ Haut-Vernet
- ☐ Nouveau logis

Acceptez-vous de participer à cette étude c'est-à-dire de répondre à ce questionnaire et de réaliser un prélèvement de sang ? / *Do you agree to participate in this study, that is to say, to answer this questionnaire and to have a blood sample taken?*

☐ oui / *yes* ☐ non / *no*

*Si non, le participant refuse → stopper l'entretien*
*If no, the participant refuses → end the interview*

Age :/ *Age:* …..

Sexe :/ *Sex:*

☐ Homme/Garçon / *Male* ☐ Femme/Fille / *Female*

Poids (en kg) :/ *Weight:* ……

Taille (en cm) :/ *Height:* …….

Antécédents médicaux :/ *Medical history:*

| | |
|---|---|
| ☐ Aucun | ☐ *None* |
| ☐ grossesse en cours | ☐ *Current pregnancy* |
| ☐ Asthme / | ☐ *Asthma* |
| ☐ Autres maladies respiratoires (bronchite chronique…) | ☐ *Other respiratory diseases* |
| ☐ Hypertension | ☐ *Hypertension* |
| ☐ maladie cardiaque (angine de poitrine, infarctus) | ☐ *Heart disease* |
| ☐ diabète traité | ☐ *Treated diabetes* |
| ☐ Cancer en cours de traitement (sauf hormonothérapie) | ☐ *Being treated for cancer (excluding hormone therapy)* |
| ☐ VIH et autres troubles de l'immunité | ☐ *HIV and other immune disorders* |
| ☐ maladies chroniques du foie | ☐ *Chronic liver disease* |
| ☐ maladies rénales chroniques | ☐ *Chronic kidney disease* |
| ☐ autre ALD, précisez : ……………… | ☐ *Other chronic disease: ……* |

Depuis le 24 février (à la fin des vacances scolaires d'hiver/semaine du Mardi Gras), avez-vous eu des symptômes que vous n'avez pas habituellement et qui ont duré au moins 3 jours ? / *Since the 24th February (at the end of the winter school holidays/Mardi Gras week), have you had any symptoms that you don't usually have and that lasted at least 3 days?*

☐ oui / *yes* ☐ non / *no*

**Si oui / *If yes:***

Quels symptôme(s) avez-vous eu ? / *What symptom(s) have you had?*

| | |
|---|---|
| ☐ Fièvre ou sensation de fièvre | ☐ *Fever or feeling feverish* |
| ☐ Mal à tête | ☐ *Headache* |
| ☐ Fatigue inhabituelle | ☐ *Unusual fatigue* |
| ☐ Courbatures / douleurs musculaires | ☐ *Body aches, muscle pain* |

☐ Toux ☐ *Cough*
☐ Difficultés respiratoires, essoufflement inhabituel ☐ *Difficulty breathing, unusual shortness of breath*
☐ Nez qui coule ☐ *Runny nose*
☐ Troubles du goût/de l'odorat ☐ *Taste/smell disorders*
☐ Nausées/vomissements ☐ *Nausea, vomiting*
☐ Diarrhée ☐ *Diarrhoea*
☐ Douleurs thoraciques, oppression ☐ *Chest pain, oppression*
☐ Si ≥80 ans : Confusion, chutes répétées ☐ *If ≥80 years: Confusion, repeated falls*

Quand ont commencé ces symptômes ? *(si plusieurs périodes : prendre les symptômes les plus proches du Covid-19 ou si impossible de différencier prendre la 1ère période)* / *When did these symptoms start? (if more than one period: record the time when the symptoms which most closely resemble those of Covid-19 started, or if it is impossible to differentiate between periods, take the first period)*

☐ avant confinement (17/03) / *Before the lockdown (17/03)*
☐ Pendant confinement (17/03) et avant 1 mai / *During the lockdown (17/03) and before the 1st of May*
☐ après le 1er mai / *After the 1st of May*

*Si après le 1er mai :* Avez-vous eu des signes au cours des 15 derniers jours ? / *If after the 1st of May: have you had any symptoms in the last fortnight?*
☐ oui / *yes* ☐ non / *no*

Ces symptômes vous ont-ils fait penser que vous aviez peut-être le coronavirus ? / *Did these symptoms lead you think that you might have COVID-19?*
☐ Non / *No* ☐ oui peut-être / *Yes, maybe* ☐ oui sûrement / *Yes, definitely*

Avez-vous consulté un professionnel de santé pour ces symptômes ? / *Did you consult a health professional for these symptoms?*
☐ oui / *yes* ☐ non / *no*

*Si oui* : qui avez-vous consulté ? / *If yes: Who did you consult?*
☐ Médecin traitant / *General practitioner*
☐ Centre covid / *Covid centre*
☐ Hôpital, urgence (sans hospitalisation) / *Hospital, emergency department (without hospitalisation)*
☐ Autre, précisez : / *other, specify:* ………

Avez-vous été hospitalisé en raison de ces symptômes ?/ *Were you hospitalized because of these symptoms?*
☐ oui / *yes* ☐ non / *no*

*Si oui* : combien de temps avez-vous été hospitalisé (en nombre de jours) ? / *If yes: how long were you hospitalised (number of days)?* : ….
Avez-vous été hospitalisé en service de réanimation ? / *Were you hospitalised in an intensive care unit?*
☐ oui / *yes* ☐ non / *no*

Avez-vous eu … ? / *Did you have a … ?*
☐ Test PCR (coton-tige) / *PCR test (swab/cotton bud)*
Si oui / *If yes* : ☐positif / *positive* ☐négatif / *negative* ☐Ne sait pas / *Don't know*
☐ Sérologie (prise de sang) / *Serology test (blood sample)*
Si oui / *If yes* : ☐positif / *positive* ☐négatif / *negative* ☐Ne sait pas / *Don't know*

□ Scanner thoracique / *Chest CT scan*

Si oui :/ *If yes:* □évocateur du Covid-19 / *suggestive* □non évocateur / *not suggestive*

□ Ne sait pas / *Don't know*

A votre connaissance depuis le 24 février, avez-vous été en contact avec une ou plusieurs personnes malades (toux ou fièvre ou test positif ou consultation pour une suspicion de coronavirus) à l'extérieur de votre logement ? / *To your knowledge, since the 24th February, have you been in contact with one or more sick people (cough or fever or positive test or consultation because of suspected COVID-19) outside your home?*

□ oui / *yes* □ non / *no* □ Ne sait pas / *Don't know*

Logement :/ *Housing:*

En ce moment, combien de personnes habitent dans le logement où vous vivez actuellement (y compris vous-même) ? / *How many people live in your current home (including you)?* ……….

Nous allons maintenant parlé du logement principal dans lequel vous viviez pendant les 2 mois du confinement. (si plusieurs endroits, prendre la plus longue durée)

*We are now going to talk about the main accommodation where you lived during the two months of lockdown (if more than one place, take the place where respondent lived longest)*

Est-ce le logement dans lequel vous habitez actuellement ? *Is that your current home?*

□ oui / *yes* □ non / *no*

*Si le participant répond non* : Vous avez passé la majorité de votre confinement dans un logement différent de votre logement actuel. Ce logement était-il dans le quartier :

*If the respondent replies 'no': You spent the majority of the lockdown in a home other than your current home. In which neighbourhood was that accommodation located?*

□ St Jacques

□ Nouveau logis

□ Haut-Vernet

□ Aucun de ces trois quartiers / *None of these three neighbourhoods*

Ce logement était :/ *This accommodation* is/*was:*

□ Un appartement / *An apartment*

□ Une maison / *A house*

□ Autre, précisez : …. / *Other, specify:…..*

Combien de pièces comportaient ce logement (hors salle de bain, toilettes, cuisine) ? / *How many rooms are/were in the accommodation (excluding bathroom, toilet and kitchen)?* : ….

Avait-t-il un espace extérieur privé (jardin, terrasse, balcon) ? / *Is/Was there a private outdoor space (garden, patio, balcony)?*

□ oui / *yes* □ non / *no*

Pendant le confinement, combien de personnes habitaient dans ce logement (y compris vous-même) ? / *During the lockdown, how many people lived in the household (including you)?* ……….

Dont combien d'enfants de moins de 12 ans :/ *Including children under 12 years:* …

Dont combien d'enfants de 12 à 17 ans :/ *Including children aged 12-17 years:* …

Dont combien d'adultes de 18 ans et plus :/ *Including adults aged 18 years and over:* …

Depuis le 24 février, combien de personnes vivant dans ce logement (autre que vous) ont été malades (toux ou fièvre ou test positif ou consultation pour une suspicion de coronavirus) ? / From *the 24th of February to the end of lockdown, how many people living in this household (excluding you) have been ill (cough, fever, positive test or consultation for suspected COVID-19)?* : …

Information et comportements face au Covid-19 / *Information and behaviour in the face of Covid-19*

Etes-vous sorti pendant le confinement pour le travail ? / *During the lockdown, did you go out for work?*

 jamais / *never*  moins d'1 fois/semaine / *less than once a week*
 1fois/semaine / *once a week*  tous les jours ou presque / *every day or almost every day*

Etes-vous sorti pendant le confinement pour les courses ? / *During the lockdown, did you go out to do the grocery shopping?*

 jamais / *never*  moins d'1 fois/semaine / *less than once a week*
 1fois/semaine / *once a week*  tous les jours ou presque / *every day or almost every day*

Etes-vous sorti pendant le confinement pour visiter la famille/des proches ? / *During the lockdown, did you go out to visit family/friends?*

 jamais / *never*  moins d'1 fois/semaine / *less than once a week*
 1fois/semaine / *once a week*  tous les jours ou presque / *every day or almost every day*

Etes-vous sorti pendant le confinement pour faire du sport/se promener ? / *During the lockdown, did you go out to play sports/for a walk?*

 jamais / *never*  moins d'1 fois/semaine / *less than once a week*
 1fois/semaine / *once a week*  tous les jours ou presque / *every day or almost every day*

Etes-vous sorti pendant le confinement pour une cérémonie ? / *During the lockdown, did you go out for a ceremony?*

 jamais / *never*  1 fois / *once*  plusieurs fois / *several times*

Etes-vous sorti pendant le confinement pour une autre raison ? / *During the lockdown, did you go out for another reason?* :  oui, précisez :/ *yes, specify:* …………………..

 jamais / *never*  moins d'1 fois/semaine / *less than once a week*
 1fois/semaine / *once a week*  tous les jours ou presque / *every day or almost every day*

Est-ce que des personnes qui n'habitaient pas dans votre logement sont venues chez vous pendant le confinement ? (Par exemple pour amener à manger ou pour des soins)
*Did people, other than people who live in the same housing as you, come to your home during the lockdown? (For example, to bring food or provide care or assistance)*

 jamais / *never*  moins d'1 fois/semaine / *less than once a week*
 1fois/semaine / *once a week*  tous les jours ou presque / *every day or almost every day*

Sur une échelle de 0 à 10, avez-vous eu des informations sur ce qu'il fallait faire pour se protéger et protéger les autres : lavage des mains, confinement, respect des distances avec d'autres personnes, port du masque ? (0=aucune information → 10 = informations complètes)
*On a scale from 0 to 10, how much information did you have on what to do to protect yourself and others: hand washing, self-isolation, social distancing, wearing a mask? (0 = no information → 10 = a great deal of information)*

0 |_______________________| 10

Avez-vous pu rester à plus d'un mètre des personnes que vous avez rencontrées à l'extérieure de votre logement pendant le confinement (par exemple : pour discuter ou dans des files d'attente) ? / *Were you able to stay more than one metre away from people you met outside your home during the lockdown ?*

□ toujours / *always* □ souvent / *often* □ rarement / *rarely* □ jamais / *never*

Vous êtes-vous lavé plus souvent les mains pendant le confinement ? / *Did you wash your hands more often during the lockdown?*

- □ non, pas plus souvent / *no, not more often*
- □ un peu plus souvent / *a little more often*
- □ beaucoup plus souvent / *much more often*
- □ Ne sait pas / *Don't know*

Sur une échelle de 0 à 10, pensez-vous vous être protégé du virus ? (0=pas du tout → 10 = complètement)

*N.B. : si demande de précision : par exemple par votre respect des gestes barrières ou par le confinement*

*On a scale from 0 to 10, do you think you protected yourself against the virus? (0 = not at all → 10 = completely)*

*N.B.: if the respondent requests clarification: for example, thanks to you respecting the preventive measures or self-isolating*

0 |______________________| 10

Si une épidémie de même nature survenait, quelle serait selon vous la mesure la plus efficace à mettre en place pour vous protéger vous et vos proches ? *In your opinion, if an epidemic of the same nature occurred in the future, what would be the most effective measure to take?*

……………………………………………………………………………………………………………………………
……………………………………………………………………………………………………………………………
